# New, fast, and precise method of COVID-19 detection in nasopharyngeal and tracheal aspirate samples combining optical spectroscopy and machine learning

**DOI:** 10.1101/2022.06.22.22276755

**Authors:** Denny M. Ceccon, Paulo Henrique R. Amaral, M. Andrade Lídia, Maria I. N. da Silva, Luis A. F. Andrade, Thais F.S. Moraes, Flavia F Bagno, Raissa P. Rocha, Daisymara Priscila de Almeida Marques, Geovane Marques Ferreira, Alice Aparecida Lourenço, Ágata Lopes Ribeiro, Jordana G. A. Coelho-dos-Reis, Flavio G. da Fonseca, J. C. Gonzalez

## Abstract

Fast, precise, and low-cost diagnostic testing to identify persons infected with SARS– CoV-2 virus is pivotal to control the global pandemic of COVID-19 that began in late 2019. The gold standard method of diagnostic recommended is the RT-qPCR test. However, this method is not universally available, and is time-consuming and requires specialized personnel, as well as sophisticated laboratories. Currently, machine learning is a useful predictive tool for biomedical applications, being able to classify data from diverse nature. Relying on the artificial intelligence learning process, spectroscopic data from nasopharyngeal swab and tracheal aspirate samples can be used to leverage characteristic patterns and nuances in healthy and infected body fluids, which allows to identify infection regardless of symptoms or any other clinical or laboratorial tests. Hence, when new measurements are performed on samples of unknown status and the corresponding data is submitted to such an algorithm, it will be possible to predict whether the source individual is infected or not. This work presents a new methodology for rapid and precise label-free diagnosing of SARS-CoV-2 infection in clinical samples, which combines spectroscopic data acquisition and analysis via artificial intelligence algorithms. Our results show an accuracy of 85% for detection of SARS-CoV-2 in nasopharyngeal swab samples collected from asymptomatic patients or with mild symptoms, as well as an accuracy of 97% in tracheal aspirate samples collected from critically ill COVID-19 patients under mechanical ventilation. Moreover, the acquisition and processing of the information is fast, simple, and cheaper than traditional approaches, suggesting this methodology as a promising tool for biomedical diagnosis vis-à-vis the emerging and re-emerging viral SARS-CoV-2 variant threats in the future.

## Introduction

After reaching the status of a pandemic, the new coronavirus variant 2 (SARS-CoV-2), the etiological agent of the 2019 coronavirus infectious disease (COVID-19), has infected more than two hundred million individuals and caused over five million deaths worldwide (https://covid19.who.int, 24/11/2021). In order to mitigate the effects of the related morbidity of COVID-19, there is a substantial need for improved population-scale testing solutions to early identify infection and thus allow adequate tracking [2]. New diagnosis techniques that are fast, accurate and low-cost will not only help the management of the current crisis, but also serve as a baseline for the development of multiplex technology that will be useful in the response to future epidemics. Presently, most diagnostic methods involve sampling and testing different fluids like nasopharyngeal cell lysate, saliva, or blood.

Infections of SARS-CoV-2 in the initial stage are currently identified using real-time quantitative polymerase chain reaction (RT-qPCR) assays, considered the Gold Standard method, which may require up to 3 days after infection for a reliable positive signal [1]. In addition, RT-qPCR tests require between 3 to 4 hours to be concluded [3] and are hardly used on a daily basis due to their elevated cost, shortages in biomarkers and key reagents.

Intermediate stage or past infections are investigated using serum-based testing methods such as enzyme linked immunosorbent assays (ELISAs), lateral flow immunoassays (LFIAs), or chemiluminescent immunoassays (CLIAs). These tests normally detect a significant and measurable concentration of immunoglobulin G (IgG) and immunoglobulin M (IgM) antibodies in blood samples. However, the build-up of such antibodies in blood is slow, thus concentrations of IgG and IgM are measurable by these methods only after two weeks of infection. [4]. Sensitive and specific serological methods are not fast, requiring 4 to 6 hours to be completed [3]. Moreover, since most infections become apparent only upon symptom onset, the current methods of testing are unlikely to identify pre-symptomatic carriers. It is estimated that as many as 50% of individuals infected with SARS-CoV-2 are asymptomatic, hampering early-stage interventions that reduce transmission [5, 6]. There is also a large number of unreported infection cases and COVID-19 related deaths [7].

In this regard, appropriate clinical samples are essential to produce reliable results for the diagnosis of infection with SARS-CoV-2. For primary diagnostic assessment for current SARS-CoV-2 infection, the Center for Disease Control and Prevention (CDC) recommends collecting and testing an upper respiratory specimen, which includes sputum, bronchoalveolar lavage and tracheal aspirate samples (CDC, 2020). Considering that the virus does not produce or poorly induces viremia, it is essential to search for the virus in the local infection milieu. As such, fast, accurate and inexpensive methods for the early detection of SARS-CoV-2 in sputum, bronchoalveolar lavage and tracheal aspirate samples in real time are urgently needed.

Emerging optical methods have been proposed for detection of virus diseases [8]. Such methods usually detect labeled samples or use laser-based expensive and complex measurement techniques [8]. However, efforts to implement fast and sensitive diagnostic approaches have emerged in response to the current health crisis, as key steps to control the pandemic as well as part of reopening strategies [9]. Although the combination of RT-qPCR and serological tests such as ELISA are ideal for an accurate diagnosis, the detection of antibodies is particularly relevant during later transmission [10]. Thus, a fast and label-free methodology for COVID-19 diagnosis during the first days after infection is desirable.

Here, we report the use of a label-free optical spectroscopic method of straightforward operation, combined with machine learning (ML) processing of the acquired spectroscopic data, as a new diagnostic method of SARS-CoV-2. Using inactivated nasopharyngeal swab samples from RT-qPCR tested individuals, as well as inactivated tracheal aspirate from intubated patients, we show that this patent pending multiplex method can be used to detect diseased individuals in less than 15 minutes, with elevated accuracy, and at a very low cost.

## Methods

### Study design and overview

We investigated whether optical spectroscopy data of nasopharyngeal swab and tracheal aspirate samples could be effectively used to detect SARS-CoV-2 infection with the aid of machine learning methods and without the use of biomarkers, in a fast and accurate way. Figure 1 shows an overview of the study process, divided in the four steps: participant recruitment, collection of nasopharyngeal swab or tracheal aspirate samples, optical spectroscopy, and machine learning modeling.

**Figure 1.**
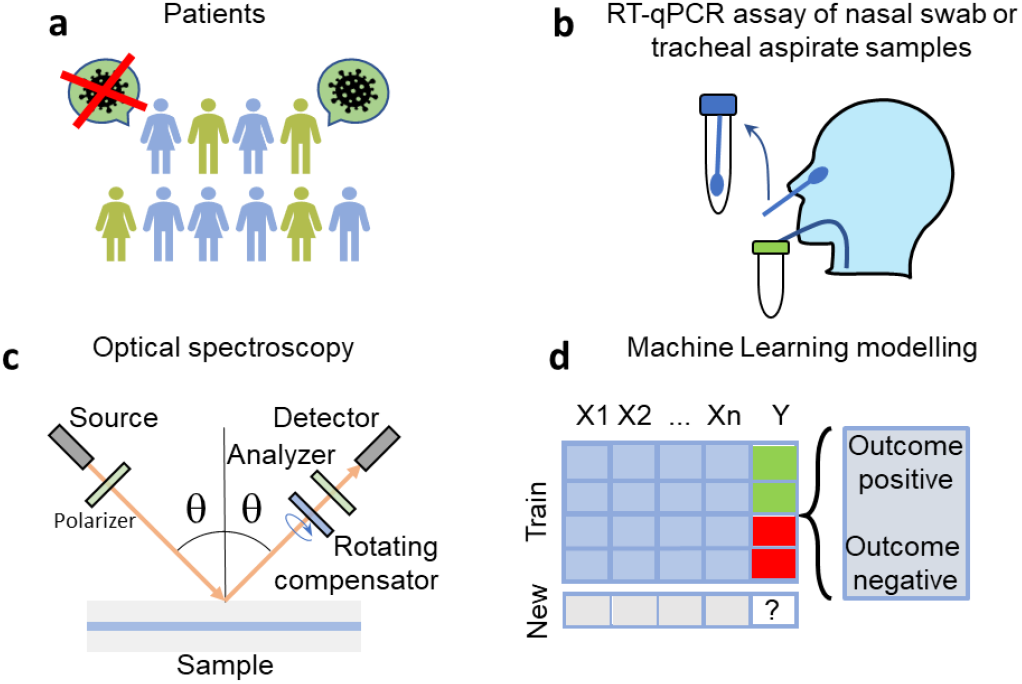
Overview of the study preocess (**a**) Enrollment. (**b**) Collection of nasopharyngeal swab or tracheal aspirate samples. (**c**) Optical spectroscopy measurements. (**d**) The algorithm was trained to predict the probability of a patient infection by COVID-19.

### Participant recruitment

The samples used in this research were collected from nasopharynx swabs of 211 patients suspected of SARS-CoV-2 infection, from asymptomatic individuals and from mildly symptomatic non-hospitalized patients. In addition, tracheal aspirate samples from 12 healthy patients and 12 critically ill COVID-19 patients, aged from 18-80 years-old, 14 males and 10 females, under mechanical ventilation at the Intensive Care Unit of Risoleta Tolentino Neves Hospital were also studied.

The use of these samples was approved by the Ethical Committee (CAAE: 32113420.6.0000.5149; 1686320.0.0000.5149) from Universidade Federal de Minas Gerais (UFMG). Sensitive information was duly anonymized. All procedures followed ethical guidelines in accordance with Brazilian national regulations.

### Collection of nasopharyngeal swab samples

Nasopharyngeal and oropharyngeal swab samples were collected from participants by inserting a rayon swab with a plastic shaft into the nostril, parallel to the palate, and gently scraped for a few seconds to absorb secretions. Another swab was shafted into the tonsils for sample collection. Next, the swabs were immediately merged into a sterile tube containing 2 mL of guanidine isothiocyanate solution. RNA extracted from all samples was tested by RT-qPCR using probes for viral and human genes. RT-qPCR was performed at the Vaccine Technology Center (CTVacinas) of the Universidade Federal de Minas Gerais to allow definitive diagnosis of SARS-CoV-2 infection. Ground truth categorization of swab samples into negative (105) versus positive (106) SARS-CoV-2 infection was based on PCR results.

### Collection of tracheal aspirate samples

Tracheal aspirate (TA) samples (2-10 mL) were collected during the early morning routine of COVID-19 patients. All patients included in the study tested positive for SARS-CoV-2 by RT-qPCR targeting the E gene. Only secretive productive patients were included in the study. Samples were aspirated into sterile tracheal secretion collectors and immediately processed in a biosecurity level 3 laboratory.

### Optical spectroscopy measurements

For the optical measurements, each nasopharyngeal swab or tracheal aspirate sample was thawed and homogenized by spinning for 1 minute, at room temperature and 1200 rpm. Next, 10 µL of the sample was deposited on a 22 mm × 22 mm glass #1½ coverslip (Corning, USA) and covered with a second coverslip. The sandwich samples were studied by ellipsometry in the 245 – 1690 nm wavelength range, with incidence angle varying from 45 to 70 degrees in 5-degree steps. The measurements were repeated in 9 different regions of approximately 3 mm × 6 mm of each slide, organized as a 3×3 rectangular mesh, in order to account for possible spatial inhomogeneities across the samples.

### Development of the machine learning model

The measured data was used to train a machine learning model to identify SARS-CoV-2 infected patients. This model was specifically trained to predict the infection status for each of the distinct positions read from the individual slides. The patient’s final diagnosis was defined by the average infection probability of all positions in the slide. An average probability below 0.5 meant a negative diagnostic, being positive otherwise.

Model design was performed in three stages: feature treatment/model type selection, training with data augmentation, and model tuning. Throughout, model quality was assessed by accuracy, precision, recall, F1 and ROC-AUC scores in a test set, determined at the patient-level. F1 was chosen as the reference metric for optimization.

In the first stage of model design, the pipeline consisted of the following sequential steps: manual variable selection, manual feature selection, scaling preprocessing, methods of outlier detection and removal, automatic feature selection, and model type selection. These steps aimed to recognize the variables, features and preprocessing procedures that would yield the best models. For manual variable selection, we considered the variables related to experimental design. The angles of incidence were tried individually (45, 50, 55, 60, 65 and 70 degrees) and combined (all). Four windows of wavelength were tried: below 380 nm, between 380 nm and 1000 nm, above 1000 nm, and the whole range (all). Due to concerns of rapid sample degradation, as well as the will to speed up the procedure in a clinical setup, three combinations of positions were tried: positions 1-3, 1-5, and 1-9 (all). For manual feature selection, we used combinations of the measured ellipsometry features: angles Ψ and Δ, depolarization, intensity, and the real and imaginary parts of the complex reflectance ratio *ρ=tan Ψ e*^*iΔ*^. The scaling step is introduced to express all measures in a comparable scale; the methods tested were MinMaxScaler, StandardScaler, QuantileTransformer and RobustScaler, as implemented by the Python package Scikit-Learn v0.24 [14]. The outlier detection methods tested were PCA, LOF, KNN, COPOD and IForest, with contamination rates in the range of 1 to 12.5%, as implemented by the Python package PyOD v0.8.7 [15]. Automatic feature selection was performed to rank the features according to their discriminative power. The methods tested in this step were ExtraTreesClassifier (both by Gini and entropy criteria), PCA, and LDA, as implemented by Scikit-Learn v0.24. After the features were ordered accordingly, we tried the top ‘n’ features from a range of 20 to 500. For model type, we tested implementations of Logistic Regression, Support Vector Machine, Gradient Boosting Classifier and Deep Neural Network (Multi-Layer Perceptron Classifier), by Scikit-Learn v0.24, and XGBoost Classifier by Python package XGBoost v1.4.0 [16].

In the second stage of model design, we tested the top performing models identified so far with a technique of data augmentation presented in [17]. The main idea of the method is to create synthetic training data by mixing the original measurements; more data tends to increase the power of generalization of the model. The synthetic data in this study was generated by averaging two measurements, making sure that only measurements from the same class and position would be mixed. The original data was also kept in the training set. The test set consisted only of original measurements.

The third and last stage consisted of tuning further the best performing models by adjusting the parameters specific to each model type. We performed an exhaustive search, tweaking some of the adjustable parameters according to each model documentation, relying once again on the data augmentation setup.

Throughout the model design protocol, models were trained with a training set and evaluated with a test set. Even though models were trained on individual positions in the slides, we made sure that the same slide would not be present in the training and test set at the same time, therefore preventing data leakage at the patient-level. These sets were generated by randomly splitting all the measurements available in a stratified fashion, reserving 20% of the patients to the test set. All metrics reported are an average of 10 such splits, produced as follows: at first, all available slides were shuffled then split into 5 folds with roughly the same size, then this process was repeated, yielding the 10 folds reported. Therefore, each of the 2 sets of 5 splits covered the whole dataset, and each patient was evaluated twice by the same model, trained with different patients each time.

## Results

### Machine learning model

Figure 2 depicts the steps of data preparation related to feature selection, prior to model implementation, for the nasopharyngeal swabs. The solid lines in panels a, b and c are the mean spectra of the physical property denoted in the y-axis, at a particular angle, measured at the wavelengths denoted in the x-axis, for all the positions in the slides. The shadow areas are the corresponding standard deviation, and the readings are separated by infection status (color coded). Each position of the slide is represented by a set of data as exemplified in Figure 2a; such a set contains readings for 9 different physical properties (Ψ, Δ, depolarization (depol), intensity, real part of *ρ*, imaginary part of *ρ*, sin(Δ), cos(Δ), tan(Ψ)), 6 different angles (45°-70°) and 674 different wavelengths, making a total of 9×6×674=36396 features available as a starting point for the development of the algorithm. After the manual selection of features, each position is represented by 198 features (Figure 2b), which contain data for one single angle (55°), one single physical property (depolarization) and a sub-range of wavelengths (above 1000 nm). Figure 2c represents the remaining features after the automatic feature selection and data scaling, where the wavelengths are ordered by their importance given by the method chosen for feature selection. At this point, 166 features remain: 166 selected wavelengths from the depolarization spectra at 55°. Figure 2d is a PCA representation of the same data shown in Figure 2c; some patients of the same status cluster together, but not all healthy and infected individuals are clearly discriminated by the data alone. The machine learning model is responsible for this final step in the classification task.

**Figure 2.**
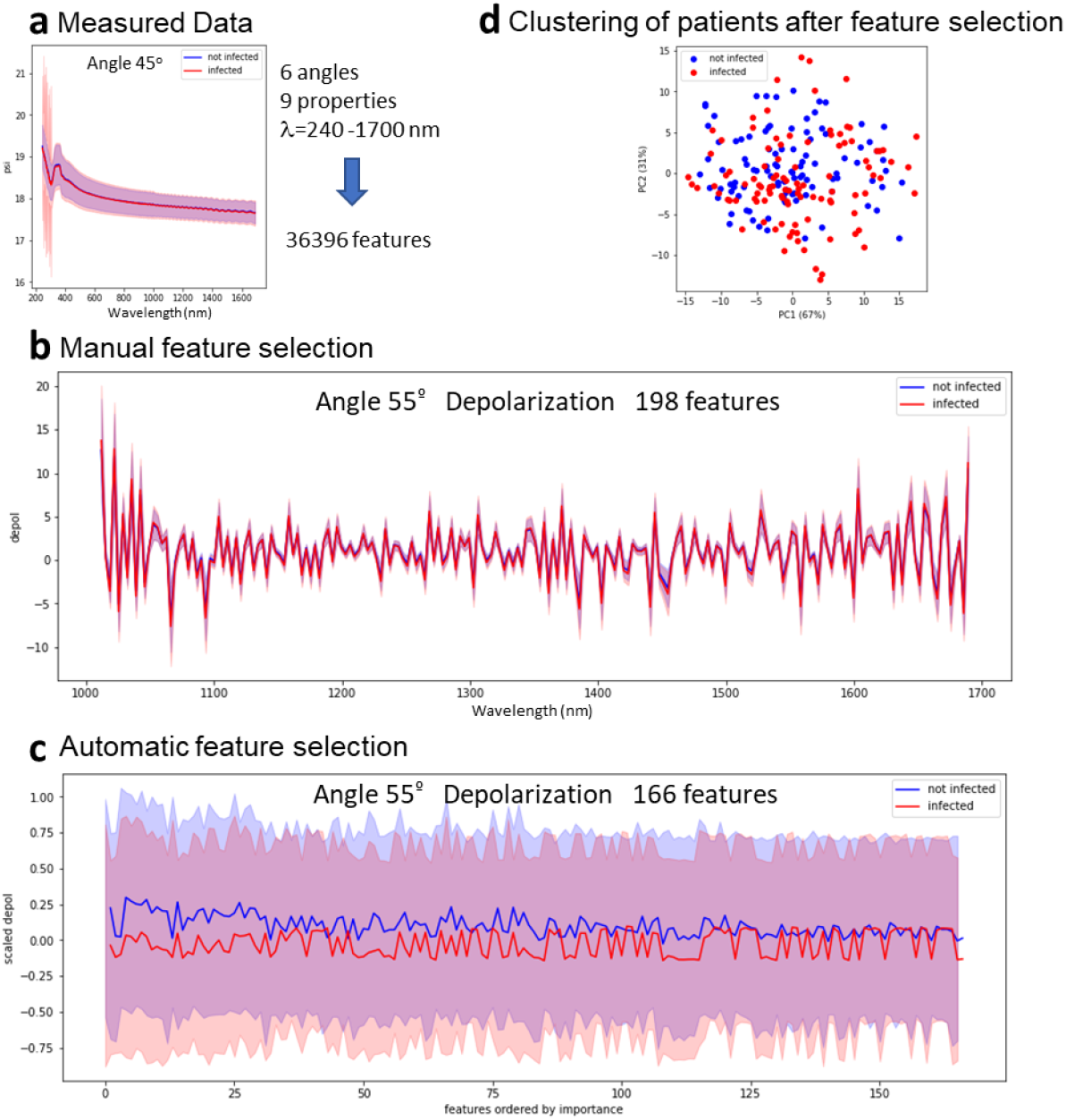
Transformation of the data along the processing steps. (**a**) Representation of the measured data. (**b**) Data after manual feature selaction. (c) Data after automatic feature selection and scaling. (**d**) PCA of the data at the end of feature selection

For these samples, the feature that delivered the best scores is depolarization, measured at an angle of 55°, at wavelengths above 1000 nm, and at all positions of a slide. They were scaled by the RobustScaler method. Outlier detection and removal was performed by the iForest method with a contamination rate of 10%. Samples from the test set were not evaluated for the presence of outliers, meaning that outliers were removed only from the training set. Automatic feature selection was guided by the ExtraTreesClassifier with Gini criterion, and 166 features were fed into the model. The model that yielded the best F1 score was an implementation of the MLPClassifier, from the Python package Scikit-Learn v0.24 [14], which is used to design neural networks. It contained two hidden layers with 100 neurons each. All layers were activated by the ReLU function. The solver used was SGD, with alpha of 1E-5, momentum of 0.95, and constant learning rate. This setup yielded a model able to diagnose patients with accuracy of 85.0% (standard deviation 6.0%), F1 of 85.9% (5.4%), precision of 79.1% (7.2%), recall of 90.4% (5.4%) and ROC-AUC of 0.900 (0.045).

In the case of the tracheal samples (Supplementary Figure 1), four features were used, Ψ, Δ, depolarization and intensity, measured at an angle of 70 degrees, at all wavelengths, at positions 1-3. The best scaling method was Robust Scaler, and outliers were removed by the KNN method with a contamination rate of 2.5%. Automatic feature selection was guided by the ExtraTreesClassification with Gini criterion, and the model performed best using 568 features. Due to the lower number of samples, the best results were achieved prior to the data augmentation phase. The best model was an implementation of the LogisticRegression classifier with standard parameters, as implemented by the Scikit-Learn v0.24 package [14]. The accuracy at the patient-level was 97.2% (standard deviation of 5.5%), F1 was 97.2% (5.7%), precision was 96.4% (7.4%), recall was 97.2% (8.3%) and ROC-AUC was 1.0.

In the configuration of 9 measured positions and only one measured angle we estimated a 7-minute interval to carry out the measurement and classification of one sample, and less than 15 minutes for the overall time of the diagnosis process of one patient, including the collection of the nasopharyngeal swab samples, preparation of the sample to be measured, the optical measurements and the AI processing of the data. The measurement and classification of the tracheal aspirate is even faster, since only 3 positions of the slide are necessary to be measured.

## Discussion

The rapid spreading of the new SARS-CoV-2 virus worldwide has shown the necessity and impact of governmental restrictions, such as lockdowns, to prevent the increase of cases and the collapse of health centers (Amaral et al., 2020). Likewise, this pandemic revealed the urgent need for fast, precise, and well-timed diagnostic systems to identify and manage the treatment of infected individuals, thus hampering the effects of COVID-Up to now, the most applied diagnostic methods encompass RT-qPCR assay at the early stage of infection, through samples collected from nasopharyngeal and oropharyngeal swabs, and ELISA at a later stage of infection by evaluating the patient’s sera (Yuan et al., 2020). Although the elevated sensitivity of current available tests, false positive and false negative results may occur depending on the time of infection and the quantity of viral load. For example, it may be challenging to find viral RNA in some samples due to the quality of transport and manipulation. Radiological methods such as chest computed tomography or thoracic radiography also have demonstrated remarkable signs of COVID-19 disease, however they cannot be used for disease screening (Adams et al., 2020).

New methodologies for massive testing are available applying LFA by different approaches, mainly using nanomaterials. Among them, an electrochemical immunoassay based on graphene electrode was functionalized with anti-spike antibodies for the rapid detection of SARS-CoV-2 virus via the spike surface protein (Mojsoska et al., 2021). Another study has proposed three-dimensional assembly of electrodes of reduced-graphene-oxide (rGO) nanoflakes immobilized with specific viral antigens integrated with a microfluidic device (Ali et al., 2020). In addition, a rapid electrochemical detection of SARS-CoV-2 antibodies using a commercially available impedance sensing platform was also proposed, which contains sensing electrodes coated with SARS-CoV-2 spike protein and exposes samples to an anti-SARS-CoV-2 monoclonal antibody (Rashed et al., 2021). However, these technologies possess some drawbacks difficult to overcome such as automation and integration of microfluidics as well as the avoidance of nonspecific biomolecules adhesion in their systems.

Plasmonic biosensors have encouraged the development of novel approaches to achieve the effective coverage of the biological receptor while confirming the affinity and specificity of targeted viral nucleic acids, proteins, or whole virus (Mauriz, 2020). Localized surface plasmon resonance (LSPR) has already been proposed to detect other viruses of medical interests such as Dengue and Zika virus (Versiani et al., 2020). Besides, other strategies using gold nanoparticles (AuNPs) serological fast-tests to identify the presence of IgM and/or IgG immunoglobulins are commercially available (Díaz-Badillo et al., 2020) and single-walled carbon nanotube (SWCNT)-based field-effect transistor (FET) semiconducting to detect the presence of SARS-CoV-2 antigens in clinical nasopharyngeal samples was assessed (Shao et al., 2021). Nevertheless, most fast-tests available have shown considerable lack of specificity (Low et al., 2021).

A more sophisticated biosensing platform was suggested by using a reverse transcription recombinase polymerase amplification (RT-RPA) coupled with clustered regularly interspaced short palindromic repeats (CRISPR-Cas12a) for the SARS-CoV-2 detection. This methodology utilizes DNA-modified gold nanoparticles (AuNPs) as a universal colorimetric readout and can specifically target the ORF1ab and N regions of the SARS-CoV-2 genome (Zhang et al., 2021). However, it is expensive and unlikely to be commercially available at large scale.

On the other hand, suggested spectroscopic techniques have demonstrated useful importance for rapid, accurate, and relatively cost-effective methods for virus detection but also for infection checking and follow-up (L. F. das C. e S. de Carvalho and Nogueira, 2020; Lukose et al., 2021). For instance, surface-enhanced Raman spectroscopy (SERS) (Saviñon-Flores et al., 2021), COVID-19 salivary Raman fingerprint (Carlomagno et al., 2021), and a superfast, reagent-free, and non-destructive approach of attenuated total reflection Fourier-transform infrared (ATR-FTIR) spectroscopy (Barauna et al., 2021) have already shown reliability for diagnostic applications.

The ability of monitoring potential virus mutations is essential, especially identifying SARS-CoV-2 variants that are known to change their RNA sequence. The use of spectroscopic techniques combined with artificial intelligence models will allow detection and probably monitor and detect any changes related to this virus (Khan and Rehman, 2020). AI has been employed in health care fields for several proposals ranging from the prediction of disease spread trajectory to the development of diagnostic and prognostic models (Syeda et al., 2021) by developing algorithms to analyze possible predictions for overall prognosis for COVID-19 patients. (Fernandes et al., 2021). Moreover, a machine-learning model that predicts a positive SARS-CoV-2 infection in a RT-PCR test based on symptoms was already established (Zoabi et al., 2021).

Despite all recent advances in diagnosis methods of SARS-CoV-2 above mentioned, there is an urgent need to develop a reagent-free, scalable, low-cost, sensitive, and specific assay for rapid detection of SARS-CoV-2 within minutes, or ideally in seconds, at the early stage of infection. Here, we have demonstrated the use of a label-free optical spectroscopy method of simple operation, combined with ML processing of the acquired raw spectroscopy data as an innovative method for SARS-CoV-2 infection detection in inactivated samples of nasopharyngeal swab and tracheal aspirate. Our methodology was validated by RT-qPCR and is applicable not only in the case of patients with mild symptoms or asymptomatic in the first stage of infection, but also to critically ill COVID-19 patients under mechanical ventilation in Intensive Care Units. Spectroscopic data from the samples, carrying information about the dielectric properties of the sample over a broad spectral range, was acquired. Software was specifically developed to manipulate the data and process them via an artificial intelligence algorithm. Both the spectroscopic technique and the software are patent pending at this moment. One of the advantages of the present method is that the samples are not labeled or processed after collection. The samples can be measured right after collection or after several weeks of storage at -20 °C. The volume of sample required for the test is relatively small, limited to 10 µL and dropped in between regular glass cover slides for measurements. Since the samples are inactivated at the moment of collection there is a very low biological risk associated with the preparation, manipulation, measurement and later discard of the slides. The simplicity and automation of the measurements and data processing procedures avoid the necessity of highly qualified personnel. These characteristics ensure the low cost of our method. In addition, a processing time of less than 15 minutes, which can be reduced with further automation of the process, and accuracy compatible with the above mentioned methods of COVID-19 diagnosis makes this solution optimal for contributing to the diagnosis of emerging infectious diseases and future pandemics of public health importance.

## Conclusion

There is a massive demand for alternative methods to detect new cases of COVID-19 as well as to investigate the epidemiology of the disease. In many countries, the importation of commercial kits poses a significant impact on their testing capacity and increases the costs for the public health system [10]. Decentralization of diagnostic testing and other technology transfer activities should be prioritized to improve accessibility in remote or isolated areas and reduce costs for the public health system [39]. Our approach demonstrates an accurate, simple, fast, label-free and cost-effective methodology for SARS-CoV-2 diagnosis.

## Data Availability

All data produced in the present study are available upon reasonable request to the authors.

## Acknowledgments

We thank all Brazilian funding agencies CAPES-PNPD scholarship program, CNPq and FAPEMIG.

